# Prevalence and factors associated with abdominal and total obesity among low-income urban population of Kiambu County, Kenya

**DOI:** 10.1101/2023.08.08.23293819

**Authors:** Grace Wambura Mbuthia, James Mwangi Ndithia, James Odhiambo Oguta, Catherine Akoth, Karani Magutah, Rosemary Kawira, Caroline Nyariki, Nickson Kimutai, Agnes Kinyua, Stephen T. McGarvey

## Abstract

Obesity is a major risk factor for most non-communicable diseases whose burden has been rising rapidly in low and middle-income countries. To develop public health interventions to address the increasing burden of overweight and obesity, estimates of the prevalence and associated factors are needed in specific populations. The study sought to determine the prevalence and factors associated with total obesity and abdominal obesity among low-income adults in Kiambu County, Kenya. This community-based cross-sectional survey involved 1656 adults residing in Kiambu County. Multistage sampling was used in the selection of participants. Data were collected by trained community health volunteers (CHVs) in their respective sub-counties using an interviewer-administered questionnaire. The CHVs also took anthropometric measurements using relevant tools and standard procedures. Descriptive statistics were used to describe the participants’ characteristics and proportions of adults with obesity. Multivariable logistic regression analyses were performed to assess the factors associated with obesity. The mean age of participants was 40.8 (±14.3) years The overall prevalence of total obesity (body mass index [BMI] ≥ 30 kg/m^2^) was 28.8% (95% CI, 26.6%-30.9%), with a higher prevalence observed among females [33.6% (95% CI, 31.1%-36.2%)] than males [12.5% (95% CI, 9.6%-16.3%)]. A third (33.3%) of the participants were overweight (25 ≤ BMI < 30 kg/m^2^). The prevalence of abdominal obesity as measured by waist-height-ratio (WHtR) was 79.8%, by waist circumference (WC) was 74.0%. Obesity/overweight by BMI was associated with female gender, increasing age, monthly income, while abdominal obesity by WHtR/WC was associated with female gender, increasing age and cigarrete smoking. In conclusion, the prevalence of total obesity and abdominal obesity was high in the population. Public health strategies focusing on weight reduction and maintenance strategies are urgently needed among low-income adults.

## Introduction

The rising burden of non-communicable diseases (NCDs) as the major cause of morbidity and mortality globally is closely paralleled by and attributable to increasing levels of obesity [1]. The worldwide prevalence of obesity nearly tripled between 1975 and 2016[2]. In 2016 over 2 billion people were overweight or obese and 70% of these people resided in the low and middle income countries(LMICs) [3]. The LMICs are undergoing a demographic transition including migration from rural to urban areas. Urban ways of life involve transitions to more physical inactivity, increased consumption of processed, nutrient dense foods, leading to increased risk of obesity and overweight. The prevalence rates of obesity are increasing more rapidly in LMICs than they are in high income countries (HICs) [4]. In LMICs, the rate of overweight and obesity is higher among females than among males and in urban areas compared to rural areas [4].

Obesity is a primary and fundamental risk factor for most cardiometabolic NCDs and contributes directly to the development of cardiovascular diseases (CVD) and CVD mortality independently of other cardiovascular risk factors [5–7]. Overweight and obesity increases the risk for type 2 diabetes, high blood pressure (BP), dyslipidemia, certain cancers, musculoskeletal disorders as well as sleep disorders[5] and, more recently, COVID 19 morbidity and mortality [8]. A combination of decline in physical activity, diets becoming sweeter, saltier, and with higher saturated fat content all which trigger the onset of obesity and non-communicable diseases Socio-economic status, age, marital status, physical inactivity and increased energy foods consumption are powerful predictors of overweight and obesity in sub-Saharan Africa [9]. In Kenya, previous studies documented high prevalence of obesity and associated adverse health outcomes [10–13]. The studies with country representative sample show a higher prevalence of overweight/ obesity among the urban population compared to the rural population in Kenya [10, 13].

Although BMI has widely been used to define severity of overweight and obesity across populations, measures of central adiposity, namely waist-to-height ratio (WHtR), waist circumference (WC) have been adopted as more accurate predictors of cardiovascular risk and have replaced BMI in several definitions for clinical diagnosis of metabolic syndrome [14–16]. These measurements are easy and affordable to collect and therefore appropriate for low-resource settings. A recent study has shown abdominal obesity as determined by WC is a CVD risk marker that is independent of BMI[5]. A WHtR cutoff of 0.5 is recommended for both sexes and in different ethnic groups and is generally accepted as a universal cutoff for abdominal obesity in adults [14]. Thus, it could be used independently being superior to both BMI and WHR, or together [17].

The purpose of this study is to describe the estimated total obesity using BMI, and abdominal obesity using WC and WHtR and the associated factors. The secondary objective will be to compare the obesity estimates from each measurement in the same study population. Previous studies in this setting [10–13] have used body mass index (BMI) as a measure of obesity at a time there is a shift to measures that focus more on abdominal obesity. The current study utilizes measures of central adiposity in addition to BMI to quantify obesity among urban low-income adults in Kenya.

## Materials and Methods

### Study Design

This was a community based cross-sectional survey. This study has been reported following internationally agreed recommendations for reporting observational studies[18].

### Study location

The study was conducted in Kiambu County, in Central Kenya.

### Study Population

Adults aged 18 years or older residing in the study area. The inclusion criteria was adult household heads aged 18 years or older, while persons with mental health disability and pregnant women were excluded. The pregnant women were excluded due to their physiological state that affects the WC measurement and BP variations during pregnancy.

### Sampling size and sampling

Multistage sampling was used to select the participants in this study. Simple random sampling was used to select Juja and Ruiru sub-counties from the 12 sub-counties in Kiambu County. Out of the 13 administrative wards in Juja and Ruiru Sub-Counties four (30% of the wards) low income wards (as characterized by the housing characteristics) were purposively sampled. Each ward is divided into 5 Community Health Units (CHU) giving a total of 20 CHUs in the four wards. Systematic sampling was used to sample 100 households from each CHU giving a total of 2000 households. Data were collected from the household heads willing to participate in the study.

### Data Collection

Trained CHVs collected data using interviewer-administered questionnaires adapted from the WHO stepwise questionnaire. Data collection took place from August-October 2022. The data collected included social-demographic characteristics and behavioral factors such alcohol use and cigarette smoking. Participants’ anthropometric measurements including height, weight and WC were taken. Weight was measured to the nearest kilogram using a calibrated bathroom scale (CAMRY Mechanical scale, BR9012, Shanghai, China), with the subject in light clothing and without shoes. Height was measured using a plastic measuring tape scaled in centimeters by asking participant to stand barefoot beside a wall on an even floor and marking the highest point of the head using a pencil and taking the measurement of the wall up to the highest point of the head. Similarly, WC was taken by placing a flexible tape measure horizontally passing it along the umbilicus and recorded at the end of normal expiration. Height and waist circumference were recorded to nearest centimeter.

### Data Analysis

Descriptive statistics were used to describe the participants’ characteristics, which were reported using frequencies, percentages, means and standard deviation. Obesity was categorized based on the World Health Organization’s BMI categories; underweight (BMI <18.5 kg/m^2^), normal weight (BMI 18.5-24.9 kg/m^2^), overweight (BMI 25-29.9 kg/m^2^) and obese (BMI ≥30 kg/m^2^) [19]. Significant WC was considered for measurements more than 90cm for males and more than 80cm in females [20] while a cut off of 0.5 for the WHtR in both gender was used to classify abdominal obesity [14]. Bivariate and multivariable binary logistic regression analyses were performed to explore the relationship between obesity and various explanatory variables. We fitted separate models for obesity as defined by different metrics (BMI, WC, WHtR). We considered a p value of 0.25 for covariates to be included in the multivariable model. We reported unadjusted and adjusted odds ratios and evaluated statistical significance at P<0.05. All the statistical analyses were performed using Stata version 17.0 (Stata Corporation, College Station, TX).

### Ethical Considerations

Ethical approval for the study was obtained from Jomo Kenyatta University of Agriculture and Technology (JKUAT) Institutional ethics committee approval number; JKU/IERC/02316/0652. Similarly, a research permit was obtained from the National Commission of Science, Technology and Innovation before commencement of the study; license number NACOSTI/P/22/19977. Participation in the study was voluntary and written informed consent was obtained from the participants before recruitment into the study. Confidentiality and anonymity of patients were guaranteed by excluding unique identifiers from the data collected from participants. We determined the WHtR by dividing the height by the waist circumference.

## Results

### Social demographic characteristics

Table 1 presents the characteristics of the sample. A total of 1,656 participants took part in the survey, indicating 83% response rate. The mean age of participants was 40.8 (SD: 14.3) years. The majority of the participants were females (77%), had secondary education (40.7%), were married (65.4%), self-employed (45.5%) and earned below 40 USD a month (41.7%).

**Table 1:** Socio demographic characteristics.

| **Variables** | **Females**<br>**n (%)** | **Males**<br>**n (%)** | **Total**<br>**n (%)** |
| --- | --- | --- | --- |
| **Age in years (N=1656)** | 40 (13.9) | 43.3 (15.1) | 40.8 (14.3) |
| 18-30 | 362 (28.4) | 85 (22.3) | 447 (26.9) |
| 31-40 | 395 (31) | 105 (27.5) | 500 (30.2) |
| 41-50 | 253 (19.7) | 75 (19.6) | 328 (19.8) |
| 51-60 | 145 (11.4) | 65 (17.0) | 210 (12.7) |
| >60 | 119 (9.3) | 52 (13.6) | 171 (10.3) |
| **Education level (N=1653)** |  |  |  |
| Never been to school | 52 (4.1) | 9 (2.3) | 61 (3.7) |
| Primary | 455 (35.8) | 104 (27.3) | 559 (33.8) |
| Secondary | 510 (40.1) | 163 (42.9) | 673 (40.7) |
| Tertiary | 255 (20.1) | 105 (27.5) | 360 (21.7) |
| **Marital status(N=1654)** |  |  |  |
| Single | 310 (24.4) | 91 (23.9) | 401 (24.3) |
| Married | 811 (63.7) | 274 (71.9) | 1085 (65.4) |
| Divorced/separated/widowed | 152 (11.9) | 16 (4.2) | 168 (10.2) |
| **Work status (N=1649)** |  |  |  |
| Employed | 148 (11.7) | 93 (24.5) | 241 (14.6) |
| Self- employed | 581 (45.8) | 168 (44.2) | 749 (45.5) |
| Homemaker | 242 (19.1) | 8 (2.1) | 250 (15.2) |
| Retired | 53 (4.2) | 22 (5.8) | 75 (4.6) |
| Student | 37 (2.9) | 14 (3.7) | 51 (3.1) |
| Casual worker | 208 (16.4) | 75 (19.7) | 283 (17.2) |
| **Income per month (USD) (N=1550)** |  |  |  |
| ≤40 | 537 (44.6) | 109 (31.5) | 646 (41.7) |
| 40-80 | 412 (34.2) | 102 (29.5) | 514 (33.2) |
| 81-120 | 112 (9.3) | 47 (13.6) | 159 (10.3) |
| >120 | 143 (11.88) | 88 (25.4) | 231 (14.9) |
| **Alcohol Use (N=1648)** |  |  |  |
| Yes | 145(11.4) | 145(38.5) | 290(17.6) |
| No | 1,126(88.6) | 232(61.5) | 1,358(82.4) |
| **Cigarette smoking(N=1,653)** |  |  |  |
| Yes | 18(1.4) | 79(20.8) | 97(5.9) |
| No | 1,255(98.6) | 301(79.2) | 1,556(94.1) |

### Prevalence of total obesity and abdominal obesity

Overall, 62.1% of the participants were overweight/obese, while only a third (562, 34%) had normal BMI. The prevalence of obesity was 28.8% (95%CI, 26.7%-30.9%). There was a higher prevalence of obesity in females compared to males (33. 6% [95% CI, 31.1%-36.2%] vs 12.5%[95% CI, 9.6%-16.3%]. The prevalence of overweight was 33.3% and was higher in females (35% [95% CI, 32.4%-37.7%]) than men (27.5% [95% CI, 23.2%-32.1%]). The WHtR estimated higher prevalence of abdominal obesity compared to WC, at 79.8% (95%CI, 77.8%-81.6%).

### Factors associated with Overweight/Obesity as measured by BMI

Results of the unadjusted logistic regression model reveal that gender, age, marital status, work status, cigarette smoking and alcohol use were all significantly associated with overweight/obesity. In the adjusted model, gender, increasing age and monthly income above 80 USD were associated with overweight/obesity (Table 3). Males had 73% lower odds of being overweight and obese (aOR=0.27; 95% CI 0.20-0.36) compared to the females. Participants in older age groups had 2-3 times higher odds of overweight/obesity compared to those aged 18-30 years. Similarly, participants earning more than 80 USD monthly increased odds of being overweight/obese.

**Table 2:** Prevalence of total obesity and abdominal obesity.

| Variables | Females<br>Mean (SD) or<br>n (%) | Males<br>Mean (SD) or<br>n (%) | Total |
| --- | --- | --- | --- |
| Weight (Kg) | 71.0 (14.7) | 68.5 (12.9) | 70.5 (14.3) |
| Height (Meters) | 1.59 (0.08) | 1.68 (0.10) | 1.61 (0.09) |
| BMI (kg/m<sup>2</sup>) | 27.9 (5.8) | 24.4 (4.7) | 27.2 (5.8) |
| Waist circumference(cm) | 93.4 (13.2) | 90.4 (11.6) | 92.7 (12.9) |
| **BMI categories (N=1652)** |  |  |  |
| Normal (BMI 18.5- 24.9) | 358 (28.2) | 204 (53.4) | 562 (34.0) |
| Underweight (BMI <18.5) | 40 (3.2) | 25 (6.5) | 65 (3.9) |
| Overweight (BMI 25-29.9) | 445 (35.0) | 105 (27.5) | 550 (**33.3**) |
| Obesity (BMI ≥30) | 427 (33.6) | 48 (12.6) | 475 (**28.8**) |
| **Waist circumference (N=1651)** |  |  |  |
| Not at risk (≤80cm for Females and ≤ 90 for male) | 224 (17.7) | 206 (53.9) | 430 (26.0) |
| At risk (WC>80cm for females and >90cm for males) | 1045 (82.3) | 176 (46.1) | 1221 (**74.0**) |
| **WHtR (N=1656)** |  |  |  |
| ≤0.5 | 209 (16.4) | 126 (33.0) | 335 (20.2) |
| >0.5 | 1065 (83.6) | 256 (67.0) | 1321 (**79.8**) |

**Kg**-Kilogram; **Kg/m<sup>2</sup>** - Kilogram per metre squared; **cm** – Centimetres; **BMI**- Body mass index; ratio; **WHtR**- Waist-Height ratio; **WC**- Waist circumference

**Table 3:** Bivariate and Multivariable Logistic Regression model for factors associated with overweight/obesity.

| Variables | cOR(95%CI) | p-value | aOR(95 % CI) | P-value |
| --- | --- | --- | --- | --- |
| **Gender** |  |  |  |  |
| Females | Ref 1 |  | Ref |  |
| Males | 0.30(0.24-0.39) | <0.001 | 0.27(0.20-0.36) | <0.001 |
| **Age** |  |  |  |  |
| 18-30 | Ref |  | Ref |  |
| 31-40 | 2.25(1.73-2.92) | <0.001 | 2.03(1.51-2.73) | <0.001 |
| 41-50 | 2.58(1.91-3.47) | <0.001 | 2.55(1.90-3.86) | <0.001 |
| 51-60 | 2.47(1.75-3.48) | <0.001 | 3.08(2.03-4.67) | <0.001 |
| >60 | 2.13(1.48-3.07) | <0.001 | 2.41(1.50-3.87) | <0.001 |
| **Education level** |  |  |  |  |
| Never been to school | Ref |  |  |  |
| Primary | 1.33(0.78-2.27) | 0.298 |  |  |
| Secondary | 1.12(0.66-1.91) | 0.672 |  |  |
| Tertiary | 1.28(0.74-2.21) | 0.381 |  |  |
| **Marital status** |  |  |  |  |
| Single | Ref |  |  |  |
| Married | 1.53(1.21-1.92) |  | 1.21(0.91-1.61) | 0.185 |
| Divorced/separated/windowed | 1.97(1.35-2.90) | <0.001 | 0.97(0.61-1.51) | 0.882 |
| **Work status** |  |  |  |  |
| Employed | Ref |  |  |  |
| Self- employed | 1.48(1.11-2.00) | 0.008 | 1.24(0.88-1.75) | 0.219 |
| Homemaker | 1.45(1.00-2.08) | 0.045 | 1.18(0.75-1.88) | 0.460 |
| Retired | 1.39(0.81-2.37) | 0.233 | 1.11(0.55-2.20) | 0.767 |
| Student | 0.29(0.15-0.57) | <0.001 | 0.57(0.27-1.21) | 0.148 |
| Casual worker | 1.22(0.86-1.74) | 0.253 | 1.21(0.80-1.82) | 0.352 |
| **Income per month (USD)** |  |  |  |  |
| ≤40 | Ref |  |  |  |
| 40-80 | 1.24(0.97-1.56) | 0.083 | 1.21(0.91-1.61) | 0.177 |
| 81-120 | 1.26(0.87-1.80) | 0.212 | 1.51(0.99-2.28) | 0.053 |
| >120 | 1.35(0.98-1.85) | 0.058 | 1.76(1.19-2.58) | 0.004 |
| **Alcohol Use** |  |  |  |  |
| No | Ref |  |  |  |
| Yes | 0.76(0.59-0.98) | 0.035 | 1.04(0.76-1.43) | 0.797 |
| **Cigarette smoking** |  |  |  |  |
| No | Ref |  |  |  |
| Yes | 0.40(0.27-0.61) | <0.001 | 0.65 (0.39-1.08) | 0.101 |

**cOR** – crude odds ratio; **aOR** –adjusted odds ratio

### Factors associated with abdominal obesity as measured by WHtR

The adjusted WHtR logistic regression model shows that gender, increasing age, education level and cigarette smoking were associated with abdominal obesity (Table 4). Males had 65% reduced odds of being overweight and obese (aOR=0.35; 95% CI 0.25-0.48) compared to the females. Higher age groups are associated with 2.8-6.8 times increased odds of abdominal obesity. Smokers had 48% reduced odds of abdominal obesity compared to non-smokers (aOR=0.52; 95% CI, 0.30-0.89).

**Table 4:**
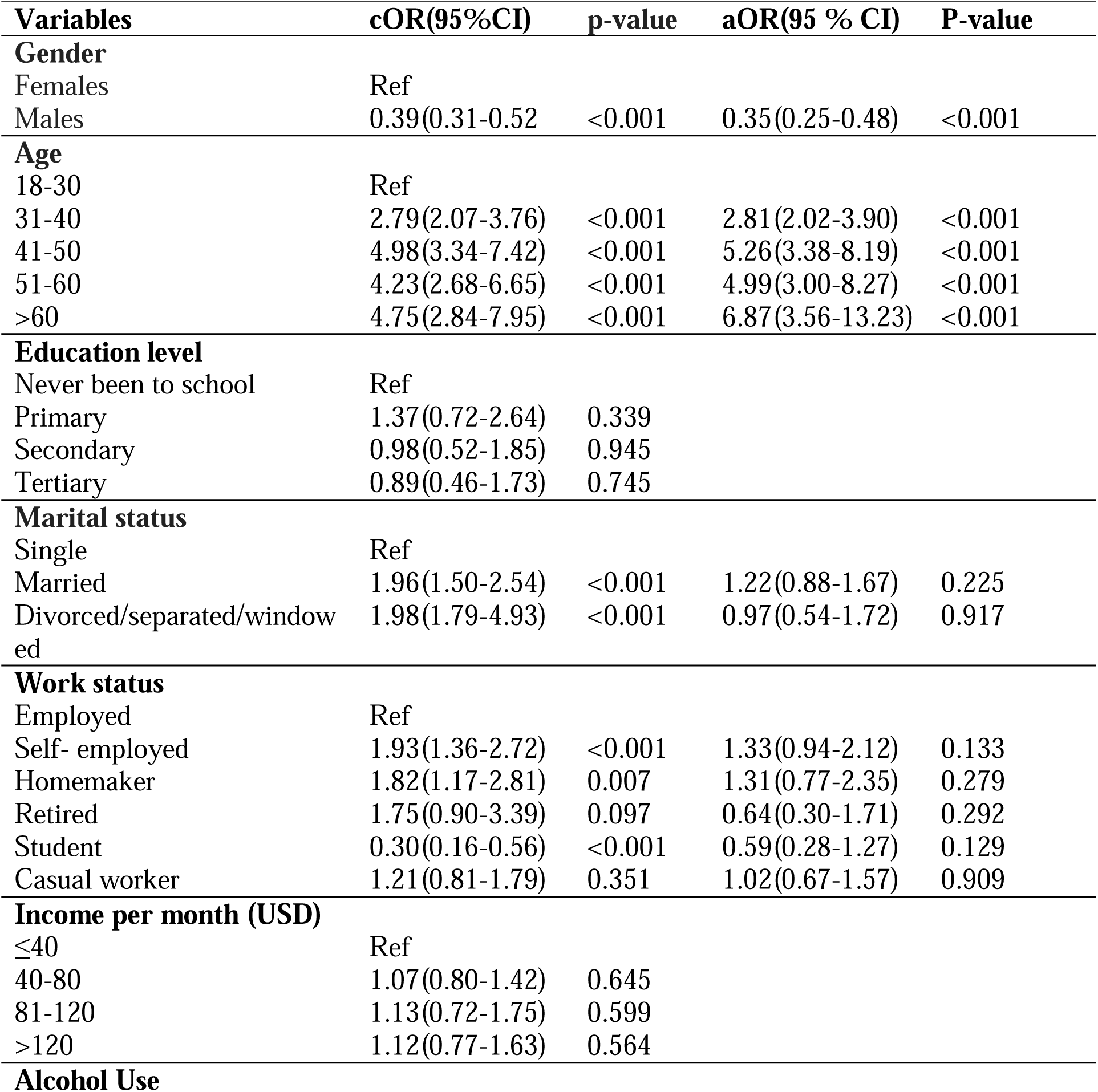
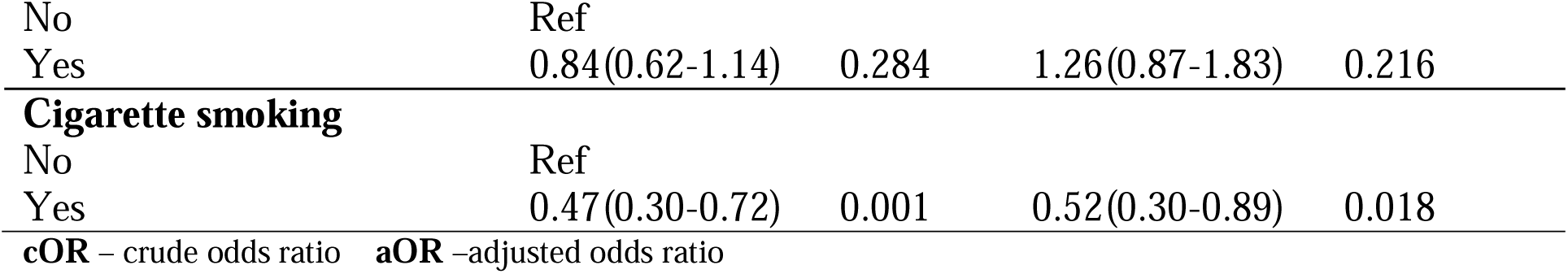
Bivariate and Multivariable Logistic Regression model for factors associated abdominal obesity by WHtR.

| Variables | cOR(95%CI) | p-value | aOR(95 % CI) | P-value |
| --- | --- | --- | --- | --- |
| **Gender** |  |  |  |  |
| Females | Ref |  |  |  |
| Males | 0.39(0.31-0.52) | <0.001 | 0.35(0.25-0.48) | <0.001 |
| **Age** |  |  |  |  |
| 18-30 | Ref |  |  |  |
| 31-40 | 2.79(2.07-3.76) | <0.001 | 2.81(2.02-3.90) | <0.001 |
| 41-50 | 4.98(3.34-7.42) | <0.001 | 5.26(3.38-8.19) | <0.001 |
| 51-60 | 4.23(2.68-6.65) | <0.001 | 4.99(3.00-8.27) | <0.001 |
| >60 | 4.75(2.84-7.95) | <0.001 | 6.87(3.56-13.23) | <0.001 |
| **Education level** |  |  |  |  |
| Never been to school | Ref |  |  |  |
| Primary | 1.37(0.72-2.64) | 0.339 |  |  |
| Secondary | 0.98(0.52-1.85) | 0.945 |  |  |
| Tertiary | 0.89(0.46-1.73) | 0.745 |  |  |
| **Marital status** |  |  |  |  |
| Single | Ref |  |  |  |
| Married | 1.96(1.50-2.54) | <0.001 | 1.22(0.88-1.67) | 0.225 |
| Divorced/separated/windowed | 1.98(1.79-4.93) | <0.001 | 0.97(0.54-1.72) | 0.917 |
| **Work status** |  |  |  |  |
| Employed | Ref |  |  |  |
| Self- employed | 1.93(1.36-2.72) | <0.001 | 1.33(0.94-2.12) | 0.133 |
| Homemaker | 1.82(1.17-2.81) | 0.007 | 1.31(0.77-2.35) | 0.279 |
| Retired | 1.75(0.90-3.39) | 0.097 | 0.64(0.30-1.71) | 0.292 |
| Student | 0.30(0.16-0.56) | <0.001 | 0.59(0.28-1.27) | 0.129 |
| Casual worker | 1.21(0.81-1.79) | 0.351 | 1.02(0.67-1.57) | 0.909 |
| **Income per month (USD)** |  |  |  |  |
| ≤40 | Ref |  |  |  |
| 40-80 | 1.07(0.80-1.42) | 0.645 |  |  |
| 81-120 | 1.13(0.72-1.75) | 0.599 |  |  |
| >120 | 1.12(0.77-1.63) | 0.564 |  |  |
| **Alcohol Use** |  |  |  |  |
| No | Ref |  |  |  |
| Yes | 0.84(0.62-1.14) | 0.284 | 1.26(0.87-1.83) | 0.216 |
| Cigarette smoking |  |  |  |  |
| No | Ref |  |  |  |
| Yes | 0.47(0.30-0.72) | 0.001 | 0.52(0.30-0.89) | 0.018 |

cOR – crude odds ratio aOR –adjusted odds ratio

### Factors associated with abdominal obesity as measured by waist circumference

Table 5 presents the results of the logistic regression analyses for the factors associated with abdominal obesity as measured by WC. The results from the adjusted model reveal that gender, age, occupation, and cigarette smoking are associated with abdominal obesity. Students had reduced odds (aOR=0.41; 95% CI 0.17-0.81) of abdominal obesity compared to being employed. Similarly, males and smokers had reduced odds of abdominal obesity compared to females and non-smokers, respectively. The odds of abdominal obesity increased with age.

**Table 5:**
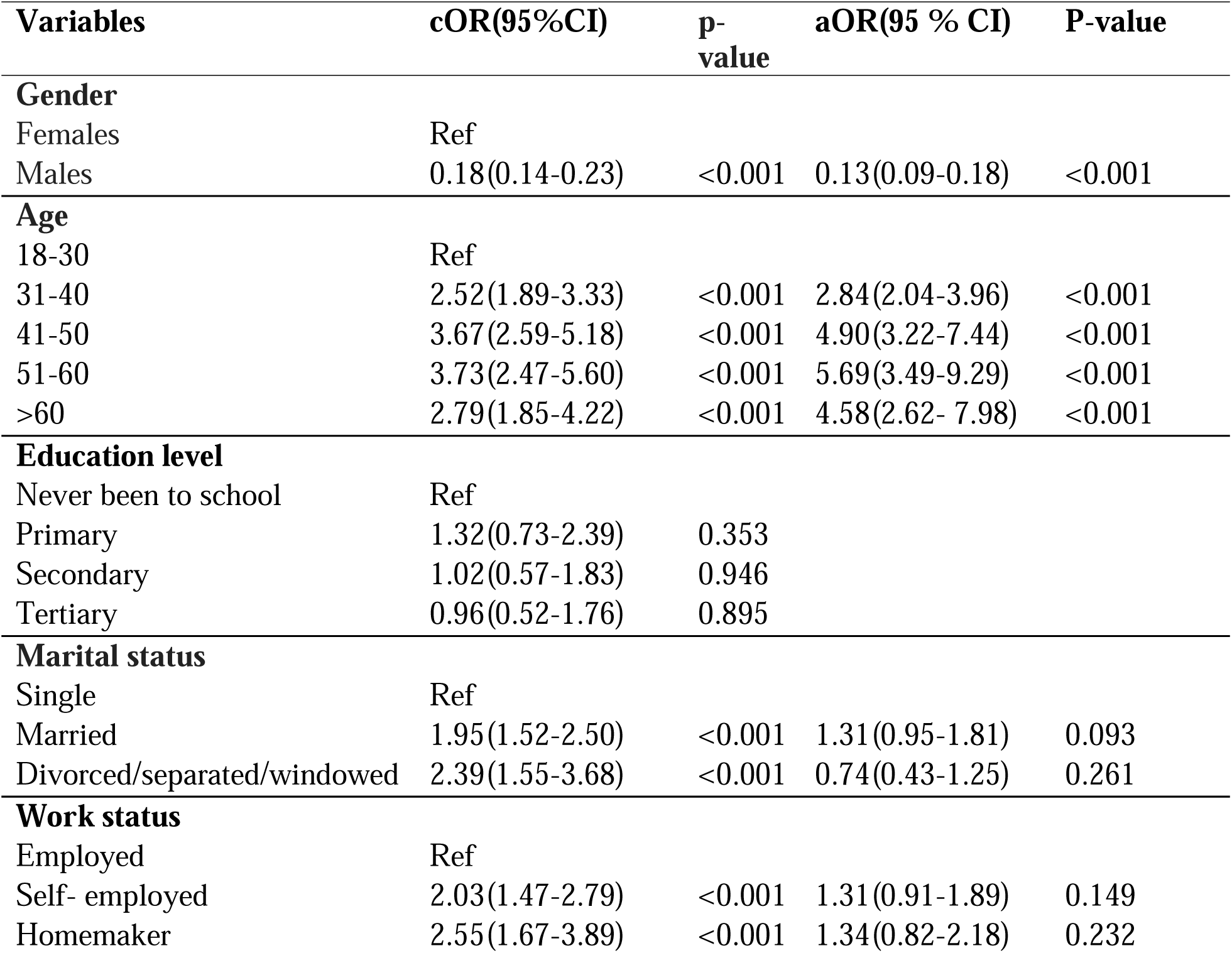
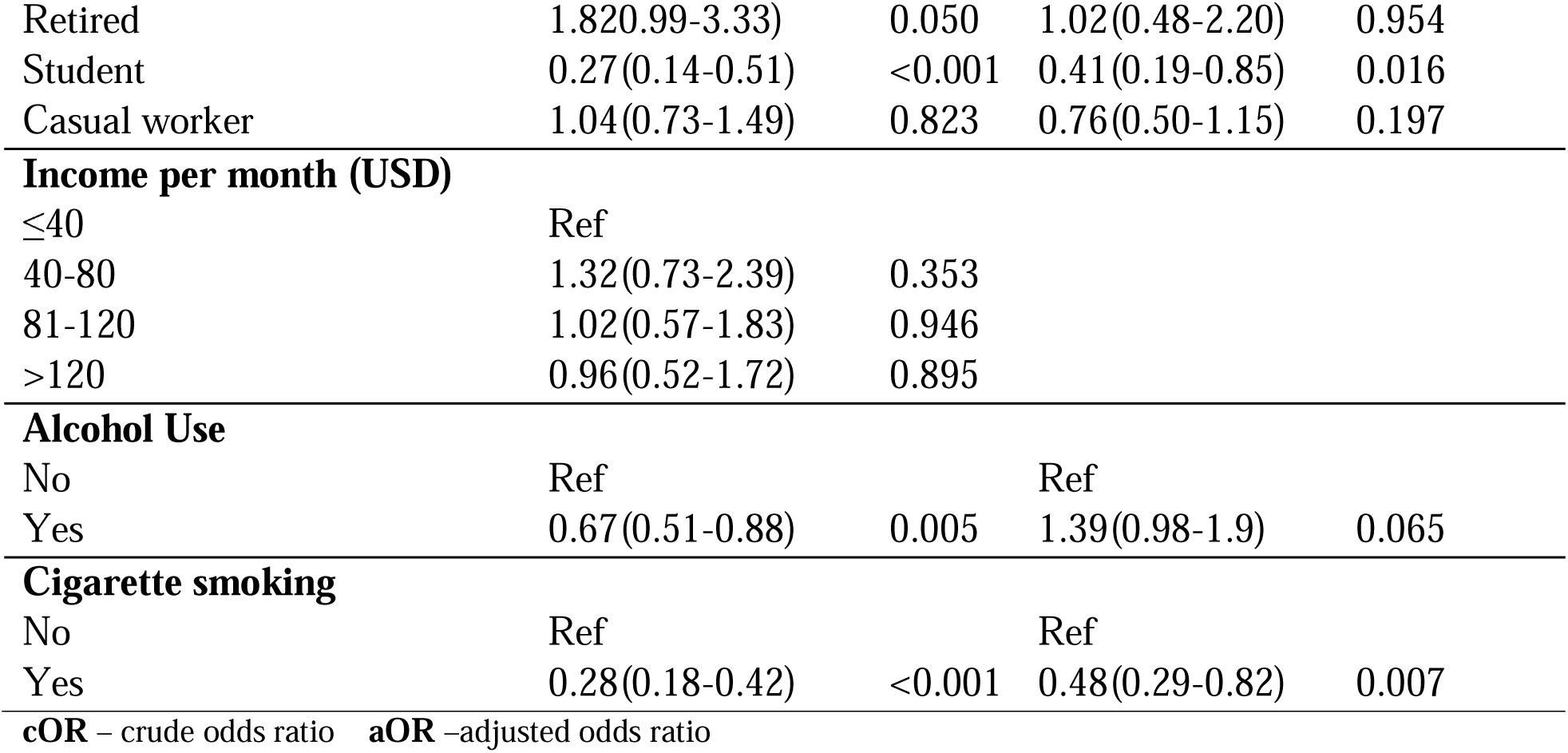
Bivariate and Multivariate Logistic Regression model for factors associated abdominal obesity by Waist Circumference.

## Discussion

Our study found a prevalence of 62.1% for overweight/obesity, and specifically 33.3% of the participants were overweight while 28.8% were obese by BMI. The prevalence was double that of a study that utilized the 2015 World Health Organization Kenya STEPwise Survey data and showed a national prevalence of overweight/obese of 31.1% [10]. Similarly, in their study, Pengpid & Peltzer reported a lower prevalence of overweight/obese of 28% in Kenya using a nationally representative community based sample [13]. The difference in the prevalence in the current study and the two study may be because the two previous studies used data collected in 2015 that included both rural and urban population. The high prevalence in this study compares to a cross-sectional study among health workers in Kenya that showed a prevalence of 58.8% for overweight/obesity[11]. The prevalence is lower than that of an urban population in a study done in Nigeria that reported a prevalence of obesity and overweight as 21% and 53.3%, respectively[21]. However, the prevalence was higher than that of a study among urban population in Norwest Ethiopia that showed a prevalence of overweight and obesity of 32% and 16% respectively[22].

The prevalence of abdominal obesity as measured by WHtR was 79.8% and WC was 74%. Our findings are similar to those of a study done in Tunisia (Traissac et al., 2015) which showed a much higher prevalence of abdominal than total obesity at 60% and 37% respectively in the same study population. On the contrary, studies measuring abdominal obesity using WC only in Southwest Iran and Southeast Ethiopia recorded a prevalence of 28.6% and 39.01% respectively, much lower than the prevalence from our study (Ghaderian et al., 2019), (Tekalegn et al., 2022). In a meta-analysis study in Ethiopia, the pooled prevalence of abdominal obesity was 37.7% (Tegegne et al., 2022). Our study measured abdominal (central) obesity using two parameters; most studies reviewed had used WC.

Comparing overall overweight/obesity prevalence using BMI and abdominal/central obesity using WHtR and WC, WHtR yielded the highest prevalence of obesity, followed by WC, with BMI, recording the lowest. Our findings are consistent with those of a study in similar settings involving adults from informal settlements in Kenya which documented a much higher prevalence of abdominal obesity by WC of 52% compared to total obesity of 20% by BMI [23]. BMI is a commonly used measure of obesity in low income settings despite its limitation in distinguishing between different body shapes/composition [24]. Previous studies among adults in urban Kenya[23] and urban Bissau in West Africa [25] have shown that individuals with normal weight may have an increased abdominal fat resulting in normal-weight central adiposity [23, 25] defined normal BMI (18.5 to 24.9 kg/m^2^) with abdominal obesity. Normal-weight central obesity is associated with higher risk of cardiovascular mortality compared to total obesity [26, 27]. Further studies to indicate the magnitude and role of normal-weight abdominal obesity in predicting cardiovascular risks in the current setting are recommended. Our findings also point to need to incorporate the other parameters in addition to BMI for detection and subsequent prevention and management of obesity.

Male participants had reduced odds of total obesity and abdominal obesity by both WC and WHtR compared to females in this study. These findings compare to those from a study with a nationally representative sample from Kenya that showed higher prevalence’s of overall and abdominal obesity in females than males[10]. Similarly, the finding is consistent with that of other studies in Ethiopia[28, 29], Malawi [30] and South Africa[31] that reported increased risk of obesity among females. Previous studies have attempted to attribute this finding to the increased pregnancies[32] and multiparous [33] nature of most females in low income settings. In addition, most men tend to engage in physically demanding and manual occupations that prevent weight gain and reduces their risk of obesity [34, 35]. Body image and societal beauty standards on females have pushed some females to desire to be heavier to feel more attractive[36]. A study in Ghana reported that 6% of females desired to be heavier, and tried to gain weight by consuming more calorie dense food[33]. Hormonal changes in females due to use of hormonal contraceptives also predispose them to obesity [37]. Moreover, postprandial lipemia, which reflects the body’s metabolism of lipids, is reportedly more among females who use oral contraceptives[38], increasing their risk for obesity. These factors may have contributed the high rates of obesity among females in this study; however, future studies to assess the extent to which the factors contribute to obesity among low-income females in Kenya are needed.

This study found that increasing age was significantly associated with higher odds of total obesity, consistent with other studies in similar settings which have reported increased risk of obesity among older adults [11, 39, 40]. Ageing leads to body composition changes, increasing fat mass and decreasing muscle mass [41] and overall metabolism, that limit movement and activity, increasing risk of obesity.

This study found that higher monthly income was associated with increased odds of being obese. Similarly, studies conducted in Ethiopia revealed that respondents who were of a better economic status were more likely to be obese compared to those of low economic status [28, 42]. The increased risk could be attributed to access to sugary and fatty diet together with a sedentary lifestyle among people with better social-economic status. However, previous studies have documented contradictory observations on the relationship between obesity and income in high and low income countries. Studies show that obesity increases with income in low income countries[43, 44] while in high-income countries social economic status is inversely associated with obesity [44, 45]. The mechanisms behind these differences is not well understood with studies suggesting that a combination of several factors to include social, cultural, psychological and biological factors play a role [43]. Further studies to investigate the causal relationship between income and obesity are needed.

Our study found smokers had reduced odds of total obesity (at bivariate) and abdominal obesity compared to non-smokers. The finding is consistent with studies done in Kenya [13], Malawi[30], Burkina Faso [46] and South-Africa [47], that showed negative associations between BMI and cigarette smoking. Similarly, previous studies showed current smokers had lower waist circumference compared to nonsmokers [48, 49]. Two possible mechanism through which cigarette smoking can lead to reduced weight /adiposity include; the effect of nicotine, which increases energy expenditure in short-term and inhibit the expected compensatory increase in caloric intake, and cigarette smoking being a behavioral alternative to eating, resulting in decreased food intake [50, 51].

The strengths of the study are in the large community based sample and the use of more than one metric to estimate obesity in the same population. However, the study has some limitations because the data on behavioral factors were self-reported, collected retrospectively and thus liable to reporting, and recall bias. Similarly, the study utilized cross-sectional design and therefore a causal relationship between the outcome and predictor variables cannot be inferred. Nevertheless, we believe our estimates represent the epidemiology of abdominal and total obesity among the low-income adults in Kiambu County and provides basis for targeted interventions to address the high obesity among urban poor and future studies.

## Conclusion

The prevalence of total obesity and abdominal obesity was high in the population. Obesity/overweight by BMI was associated with female gender, increasing age, monthly income, while abdominal obesity by WHtR/WC was associated with female gender, increasing age and cigarrete smoking. Public health strategies focusing on weight reduction and maintenance strategies are urgently needed among low-income adults.

## Conflicts of interest

The authors declare that they have no conflicts of interest.

## Funding statement

This research was supported by the Consortium for Advanced Research Training in Africa (CARTA). CARTA is jointly led by the African Population and Health Research Center and the University of the Witwatersrand and funded by the Carnegie Corporation of New York (Grant No. G-19-57145), Sida (Grant No:54100113), Uppsala Monitoring Center, Norwegian Agency for Development Cooperation (Norad), and by the Wellcome Trust [reference no. 107768/Z/15/Z] and the UK Foreign, Commonwealth & Development Office, with support from the Developing Excellence in Leadership, Training and Science in Africa (DELTAS Africa) programme. The statements made and views expressed are solely the responsibility of the authors.

## Supporting information

Data set

## Data Availability

All relevant data are within the manuscript and its Supporting Information files.

## Acknowledgement

Our special thanks go to the participants in this study for their invaluable contributions to this study. We thank the community health workers in Juja and Ruiru who conducted the data collection exercise. We also thank the Sub-county management teams and specifically the community strategy coordinators in the two sub-counties; Ms. Ann Wamaitha Mwangi and Ms. Njoki Mwaura, as well as the Community health assistants in the participating wards for their immense support during the data collection exercise.

## Notes

### Competing Interest Statement

The authors have declared no competing interest.

### Author Declarations

Ethical approval for the study was obtained from Jomo Kenyatta University of Agriculture and Technology (JKUAT) Institutional ethics committee approval number; JKU/IERC/02316/0652.

### Summary of Updates

We have revised the study aim in the abstract to include prevalence and factors associated. We also revised the background for clarity and removed statements that were repetitive. The results section was substantially revised after reanalysing the data to ensure only covariates that had a p value of 0.25 were fitted in the final multivariate model. The discussion was edited to suit the revised analysis.

